# In vivo inhibition of miR-125b modulates monocyte trafficking through the CCR7 receptor and attenuates atherosclerosis

**DOI:** 10.1101/2024.03.25.24304874

**Authors:** Adrian Mallén, Cristian Varela, Noemí Rotllan, Valentina Paloschi, Lars Mäegdefessel, Joan Carles Escolà, Josep Maria Aran, Estanis Navarro, Miguel Hueso

## Abstract

**Background:** Atherosclerosis (ATH) is a chronic systemic inflammatory disease affecting the vessel wall, wherein regulating non-coding RNAs play a crucial role. We previously demonstrated that miR-125b is upregulated in ATH and is a main regulator of cholesterol metabolism in macrophages. Herein we hypothesized that inhibiting miR-125b may attenuate ATH.

**Methods and results:** In the ApoE^-/-^ mice model fed with a high fat diet for 14 weeks, we inhibited miR-125b using an antagomiR over a 4-week period. We observed a significant reduction in plaque size, accompanied by diminished infiltration of F4/80 macrophages and attenuation of NF-κB^+^ activation within plaques. We explored the mechanism using a Vas-on-Chip adhesion assay using Human Aortic Endothelial Cells (HAoEC) stimulated with TNFα. We observed an impairment in the trafficking of miR-125b transfected THP-1 monocytes, accompanied by the downregulation of the CD11b/CD18 integrin and the CCR7 receptor. Furthermore, we demonstrated a direct regulation of the CCR7 receptor by miR-125b using a reporter plasmid construct (p_CCR7.WT) containing the 3’UTR region of CCR7 gene fused with a luciferase coding sequence. In addition, miR-125b transfected monocytes inhibited CCR7 cell migration induced by the CCL21 ligand but did not affect migration induced by others ligands such as MCP1. Finally, we confirmed the downregulation of CCR7 in coronary plaques in both ApoE^-/-^ mice and patients with coronary artery disease.

**Conclusions:** Inhibiting miR-125b offers a novel therapeutic approach for ameliorating ATH that results in a reduction of macrophage content and plaque lesion size. This improvement occurs through the enhancement of monocyte trafficking via CCR7 that facilitates the exit of foam cells from the plaque.

**CLINICAL PERSPECTIVE:** *What is New?:*

- We found evidences of a new therapeutic approach for atherosclerosis, in which miR-125b inhibition reduces macrophage content and plaque size.
- We described the molecular mechanism underlying miR-125b, which involves regulating of monocyte trafficking to plaques and the downregulation of the chemokine receptor CCR7. CCR7 plays a crucial role in facilitating the egress of macrophages and foam cells from plaques, and its downregulation contribute to progression of ATH.
- The results have been validated in a cohort of patients with coronary artery disease, where CCR7 expression was reduced in plaques.

*What are the clinical implications?:*

- We highlight the pivotal role of monocyte trafficking in the inflammatory mechanism of atherosclerosis. Managing miR-125b/CCR7 signaling may improve the resolution of ATH promoting the exit of foam cells from plaque.
- Inhibition of miR-125b in plaque macrophages represents a novel and promising therapeutic approach for cardiovascular disease.

## 1. Introduction

Atherosclerosis (ATH) is a chronic systemic inflammatory disease of the vessel wall caused by a combination of multiple factors which ranges from genomics and epigenetic modifications related to environmental conditions which eventually drives to acute cardiovascular events worldwide^1^. ATH progression is characterized by the accumulation of oxidized low-density lipoprotein (oxLDL) in the subendothelial wall, causing an inflammatory cell recruitment and accumulation, arterial intimal thickening and extracellular fibrous deposition which lead to plaque development and rupture^2^. It is also well noted that shear stress (SS) is another component that affects the initiation and development of ATH since endothelial cells (ECs) integrity is altered by these mechanical forces. High shear stress (HSS) conditions in proximal regions also induces the formation of plaque vulnerability through vascular outward remodeling, angiogenesis and vascular smooth muscle cells (VSMC) apoptosis^3,4^. In this manner, the pathophysiology underlying this cardiovascular disease include heterogeneous and complex molecular mechanisms involving mainly ECs dysfunction, monocytes recruitment and macrophage foam cell formation and VSMC migration^2,5^. Importantly, proinflammatory chemokines are released when loss of vascular homeostasis occurred. The Chemokine C-C Ligand 2 and 5 (CCL2, CCL5), Interleukin 1 beta and 6 (IL-1β, IL-6) and tumor necrosis factor alpha (TNFα) are primary produced and triggers monocyte attraction, tethering, firm adhesion and transmigration into inflamed endothelium through monocytic selectin (Cluster of differentiation molecule 62L; CD62L), integrins (cluster of differentiation molecule 11B and 18; CD11b, CD18), and chemokine receptors (Chemokine C-C Receptors 2 and 5; CCR2, CCR5)^6,7^. Within inflamed region, several macrophage receptors contribute to atherosclerosis development and foam cell formation such as oxLDL uptake receptors (e.g., scavenger receptors including CD36 and SR-AI, and lectin-like oxLDL receptor-1 (LOX-1)) and cholesterol transport receptors (e.g., ATP-binding cassette transporters A1 and G1, ABCA1 and ABCG1, and scavenger receptor SR-BI) among others^2,8,9^. Moreover, chemokine receptors such as CCR7 have been logically proposed to participate in the egress of macrophages and foam cells from the atherosclerotic lesions to lymphatic vessels via its potent chemoattractant ligands CCL19 and CCL21^8,10^.

In that regard, the focus of research on the genomic basis of ATH for discovering novel therapies and developing a more personalized medicine, has revealed in the last years that a novel group of non-coding RNAs can regulate the function of endothelial, macrophage and VSMCs and they are implicated in the onset and progression of atherosclerotic lesions. MicroRNAs are a highly conserved family of small non-coding RNAs which modulate at post-transcriptional level the gene function by base-pair binding to the target mRNA^11^. Several studies have demonstrated that expression of microRNA or “atheromiRs” within diverse type of arterial cells is altered in atherosclerotic lesions^12^. Some miRNAs, including miR-221, miR-30, miR-134a, miR-126, miR-217 and miR-223, have an important role in endothelial function, senescence and apoptosis^11,13–15^. The VSCM function, plasticity, fate determination and calcification are also regulated by a large number of miRNAs such as miR-21, miR-29, miR-125a, miR-125b, miR-143 and miR-214 among others^11,16,17^. Lipid retention, cholesterol metabolism and macrophage-mediated local inflammation are also regulated by a huge number of miRNAs, including miR-125a, miR-125b, miR-122, miR-33, miR-17^11,18,19^. Furthermore, shear stress imposed on ECs stimulates changes and also contributes to several miRNAs regulation (e.g., miR-10a, miR-23b, miR-92a, miR-101)^11,20^.

Concretely, miR-125b play a double-edge role in ATH formation and prevention including the anti-atherogenic paper in ECs but also the contribution to pro-inflammatory phenotype into ECs leading to a reduction of endothelial permeability increasing the risk of ATH events^21–23^. MiR-125b also produced a pathogenic effect on VSMC as showed in Villeneuve et al. by enhancing the production of IL-6 and MCP-1^24^, but contrary effects were found in calcified aortas with reduced expression of miR-125b in VSMCs^21,24,25^. Apart from playing an important role in ECs and VSMCs of vascular diseases, miR-125b has also been found to act as a key regulator in monocyte and macrophage cells involved in atherosclerotic lesions^21,26^. Our group identified that miR-125b is highly overexpressed in atherosclerotic plaques of abdominal aortas from coronary artery disease (CAD) deceased patients, but also in activated mice macrophage under the CD-40/Nf-Kb signaling axis^27^. More in depth we also discovered that miR-125b is a crucial regulator of lipoprotein metabolism in macrophages by targeting the 3’UTR SCARB gene which leads to a decrease in SR-BI-mediated macrophage cholesterol efflux to HDL^28^.

These data suggest that inhibition of miR-125b may result in an improvement in atherosclerotic lesions. Using a combination of in vivo and in vitro assays, we demonstrate herein, that miR-125b blockade upregulate CCR7 expression, reduce ATH lesions and macrophage infiltration in the plaque. Accordingly, we notably found that aortic plaques from human CAD patients with an increase in miR-125b, also showed a considerable downregulation in CCR7 expression. These data reinforced the relevance of novel therapies against ATH based on microRNAs.

## 2. Materials and Methods

### 2.1 Animals studies

*Apolipoprotein E knock out (ApoE*^-/-^) mice on C57BL/6 background (24 females and 24 males; Jackson Laboratories, Bar Harbor, USA) with 8-week-old were used for this experiment. To accelerate atherosclerosis, ApoE^-/-^ mice were fed from their 8-week-old with a “high-fat rodent diet” (HFD) *ad libitum* that contained 1.25% cholesterol and provided 40% of the energy as fat (D12108CI; Research Diets Inc, New Brunswick, NJ, USA) during 10 weeks. Mice from 18-week old were randomized into four groups: PBS group; antagomiR-Scramble (Anti-Sc) group; antagomiR-125b (Anti-125b) group and mimic miR-125b (miR-125b) group, having 50% (n=6) female and 50% (n=6) male in each group. The mice received intraperitoneal injections of PBS or 15mg/kg body weight Anti-Sc, Anti-125b or miR-125b twice a week for four weeks. After 4 weeks of treatments, animals were euthanized by inhalation of <5% isoflurane and cardiac puncture. The study design is summarized in Figure 1. Nucleic acid sequences of oligonucleotides used in this experiment and their modifications are described in Supplementary Table 1. All the experimental procedures were approved by the Ethics Committee for Animal Research of UB-Bellvitge and performed in accordance with the European legislation on Laboratory Animal Experiments.

**Figure 1.**
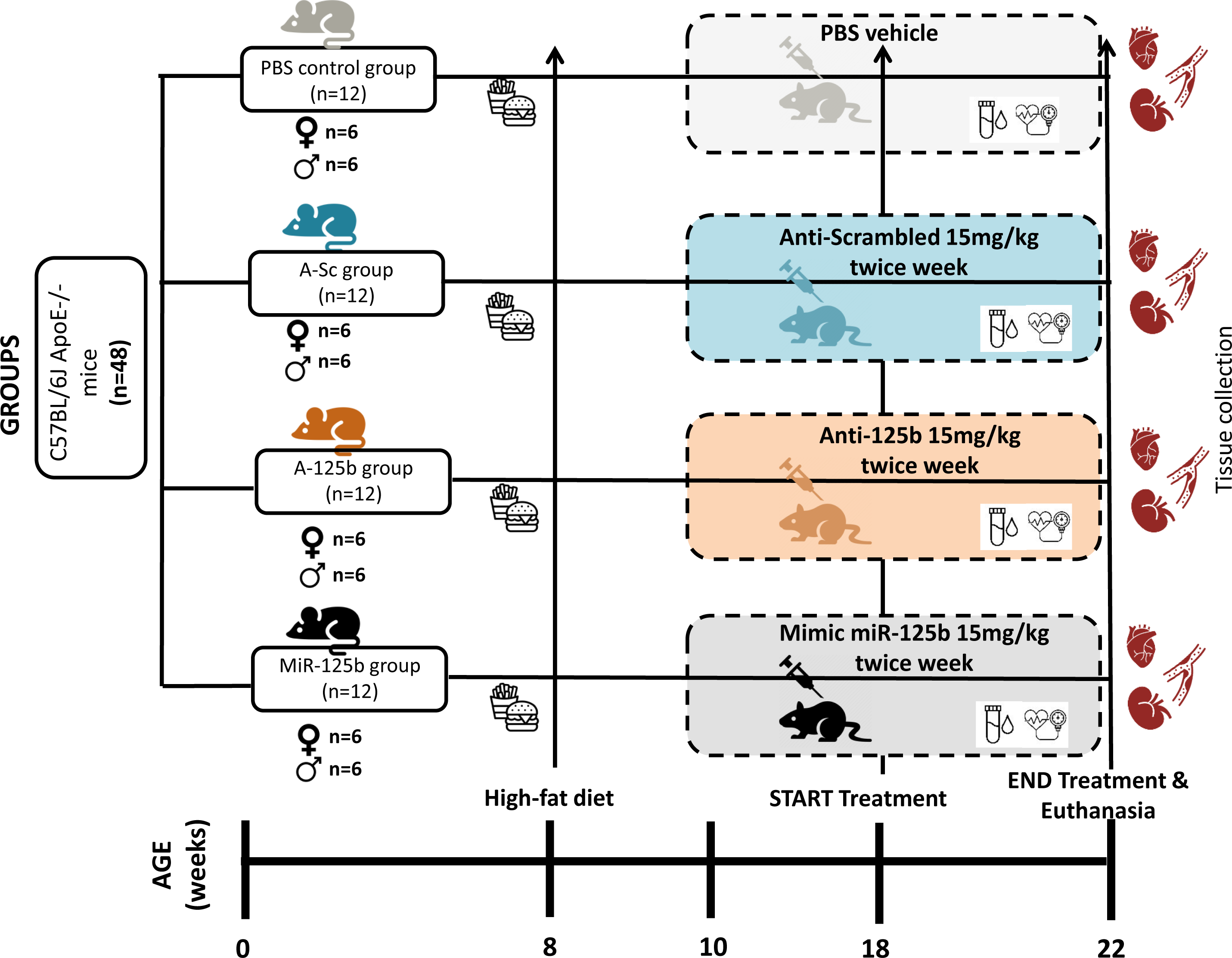
Diagram of the experimental study design showing mice treatment groups, chronogram in weeks, interventions and sample collection. A total number of 48 ApoE^-/-^ 8-week-old mice were fed with a High fat diet for 10 weeks in order to accelerate atherosclerotic plaques. Then, animals were randomized into four groups (half male and half female) depending on the treatment application. The mice received intraperitoneal injections of PBS or 15mg/kg body weight antagomiR-Scramble (n=12), antagomiR-125b (n=12) or miR-125b mimic (n=12) twice a week for four weeks. At the end of the treatment, urine samples were collected in metabolic cages and blood pressure was analyzed by non-invasive tail-cuff method. Animals were euthanized at 22 weeks of age by cardiac puncture and serums were collected. Blood was collected at the moment of sacrifice and the aortic tree, heart and kidneys were perfused with PBS and were subsequently fixed in 4% paraformaldehyde during 12 h and in 20% sucrose for 24 h. Then, heart and kidneys were embedded in optimal cutting temperature (OCT) medium, frozen immediately under liquid nitrogen and stored at -80°C for further analyses.

### 2.2 Blood pressure analysis and biochemistry parameters analysis

Blood pressure (BP) was measured by using a non-invasively tail-cuff sphygmomanometer (BP-200 Blood Pressure Analysis System NIBP LE5001, Panlab Harvard Apparatus, Spain). Urine samples were collected using metabolic chambers and plasma was also collected at the end of the study Biochemical parameters were measured at the Clinical Veterinary Biochemistry Service of the Universitat Autònoma of Barcelona (UAB) using colorimetric assays and the Beckman Coulter AU480 Analyzer (Beckman Coulter, California, USA).

### 2.3 RNA isolation and quantitative real-time PCR

Total RNA from cells was isolated following the manufacturer’s protocol miRNeasy Micro Kit (217084, Qiagen, Hilden, Germany). One microgram of total extracted RNA was reverse-transcribed using the High Capacity cDNA Reverse Transcription Kit (4368814, ThermoFisher Scientifics, MA USA) according to the manufacturer’s protocol. Next, quantitative Real-time PCR amplification was carried out in duplicates using the TaqMan Fast Advanced Master Mix (4444557, Applied Biosystems, Foster City, CA) with their specific TaqMan probes (Applied Biosystems, Foster City, CA) listed in Supplementary Table II. The amplifications were performed on a QuantStudio Pro 7 thermocycler (ThermoFisher Scientifics, MA USA) and the relative mRNA expression levels were quantified using the ΔΔCt method normalizing to GAPDH or RPLP0 internal control genes.

### 2.4 MicroRNA quantification

A total amount of 10 ng extracted RNA was used for reverse transcription and cDNA production through TaqMan MicroRNA Reverse Transcription Kit (4366596, Applied Biosystems, Foster City, CA) and specific microRNA primers for human and mouse listed in Supplementary Table III. The qPCR was performed in the same way as for the mRNA in the QuantStudio Pro 7 but miRNA expression levels were normalized to the U6 snRNA control gene.

### 2.5 Histology and Atherosclerotic Lesions Analysis

As previously described hearts were perfused, fixed, OCT-embedded and serially sectioned (10 µm) starting in the ascending aorta of the aortic root up to the aortic sinus^29^. The serial slides were stained with Masson’s trichrome for lesion area quantification, or with Oil-Red-O (ORO) and counterstained with hematoxylin in order to analyze aortic root neutral lipid content all together in the same anatomic location (170 µm after the appearance of the 3^rd^ aortic valve). Whole aortas were removed, dissected and fixed in 4% paraformaldehyde for 12 h and in 20% sucrose for 24 h. The atherosclerotic plaque was calculated by normalizing the plaque lesion area in mm^2^ (stained by Masson’s) with the total area of the section and the neutral lipid content was calculated by the percentage of positive ORO normalized also by total area of the section. Then, thoracic aorta and aortic arch were cut longitudinally, open and pinned out in a black surface showing the “en-face” area for the subsequent lipid-staining with ORO. Images of whole aortas were captured with a digital camera mounted on a dissection microscope and aortic roots images were acquired with the Nikon Epifluorescence microscope (E800) with 4X, 10X and 20X magnification and imported into ProgRes Captures (Jenoptik, Jena, Germany). En-face plaque lesion area (stained in red with ORO) was quantified morphometrically using ImageJ software and normalized by the total area of the thoracic aorta.

### 2.6 Immunohistochemistry of frozen aortic roots

Aortic roots sections immediately adjacent to ORO-stained slides were stained with Rat anti-F4/80 to study the number of infiltrated macrophages, with Rabbit anti-Nf-κB to evaluate the inflammation in the plaque and with Rabbit anti-CCR7 in order to study the number of cells that express CCR7 receptor. Corresponding secondary antibodies were applied before the DAB development and were finally counterstained with Harris hematoxylin, covering with CV Mount mounting medium (Leica, #14046430011). Dilution, brand and catalog from primary and secondary antibodies are listed in Supplementary Table IV. The relative number of positive cells was calculated by counting the positive cells inside 5 separated areas (corresponding to a total of 100 cells per area) within the aortic lesion of the same section and the mean average of 5 areas was normalized by total number of cells in each mouse.

### 2.7 Dual luciferase assays

Murine RAW264.2 were seeded in a 96-well opaque plates at 2×10^5^ cells/well (#6005660; Perkin Elmer, Massachusetts, USA) in order to identify whether the CCR7 3’UTR region is regulated by miR-125b. The cells were then transfected with 100 ng of pMirTarget reporter constructs (PS100062, Origene Technologies, Maryland, USA): the p_CCR7.WT plasmid with the wild type 3′UTR region of CCR7 gene (#SC211596; Origene pCCR7–3′UTR CCR7 (NM_001838) Human 3′UTR Clone) and p_CCR7.Mut with the mutated miR-125b predicted binding site from CCR7-3′UTR (#SC2211596_CW306881; Origene pCCR7–3′UTR CCR7 (NM_001838) alone as reference controls or together with 40 nM mirVana hsa-miR-125b-5p miRNa mimic (#4464066-MC10148; Thermofisher Scientifics, Massachusetts USA) using 0.3 μL TransIT-X2 (#6004 MirusBio, Wisconsin USA) according to the manufacturer’s protocol. After 48h post-transfection, the luciferase activity was measured using Luciferase Assay System Kit (#E4530; Promega, Wisconsin USA) as previously described^28^. Luminescence luciferase values were normalized to RFP values.

### 2.8 Cell Culture and monocyte transfection

RAW264.2 cells were cultured in DMEM medium (Gibco, #41966-029, Grand Island, USA) supplemented with 10 % FBS, L-Glutamine 1 % and Penicillin-Streptomycin 1 %. THP-1 monocyte cells were cultured in RPMI1640 medium (Gibco, #11875093, Grand Island, USA) supplemented with 20 % FBS, L-Glu 1% and P/S 1% in a concentration of 5×10^5^ cells/mL. Human aortic endothelial primary cells HAoEC (Cell Applications, #304-05a, San Diego, USA) were cultured with Endothelial Cell medium (Pelobiotech, #PB-MH-100-2190, Planegg, Germany). All medium were replaced every 2 days and cells were maintained with 5 % CO_2_ and 37°C.

A total of 1.4×10^6^ cell/mL THP-1 cells were seeded in a 6-well plate and were transfected with 40 nM of hsa-miR-125b mimic (Thermofisher, #4464066-MC10148) or 40 nM Scramble (Thermofisher, #4464058) using Lipofectamine RNAiMax (Thermofisher, #13778150) reagent with OptiMEM medium (Gibco, #31985070) for 48 h. Extra monocyte transfection supplements were used: 1 % sodium pyruvate (Gibco, #11360070), 1 % non-essential amino acids (Gibco, #11140050) and 50 µM β-Mercaptoethanol (Gibco, #11528926) were added into each well in order to improve transfection efficiency.

### 2.9 Flow adhesion assay in Vas-on-Chip

HAoEC were seeded in Leuer µ-Slider 0.6 channels (IBIDI, #80176, Gräfelfing, Germany) pre-treated with 0.1 mg/mL Collagen type I rat tail (Corning, #354236, NY, USA) for 2 h at 37 °C in a concentration of 1.6×10^6^ cell/mL until they were attached. Then, HAoEC were stimulated with 5 ng/mL TNF*α* (Thermofisher, #PHC3015) for 12 h. Subsequently, THP-1 transfected cells were labeled with 0.5 μM Cell-Trace Calcein AM (Thermofisher, #C34851), washed with PBS and resuspended at a concentration of 0.1×10^6^ cell/mL in Flow buffer (50 mL OptiMEM, 2 % FBS, 1 % P/S). Labeled monocytes were perfused over the HAoEC for 4 h under static or flow conditions (Shear stress of 0 or 12 dyn/cm^2^) using IBIDI pump system (IBIDI, #10902, Gräfelfing, Germany) and IBIDI perfusion set kit (IBIDI, #10962, Gräfelfing, Germany) with a flow rate of 23 mL/min. After flow assay, µ-Slides were washed with PBS and images of the adherent labeled monocytes were taken (Leica Thunder Microscope, Leica Microsystems, Switzerland). After imaging, the remaining adherent cells were lysed with TriZol and total RNA was extracted for RT-qPCR analysis. For each experiment, 8 random areas were selected and the percentage of THP-1 attached cells was quantified as the average count cells per area and was analyzed by ImageJ v1.46.

### 2.10 Transwell migration assay

CytoSelect^TM^ 96-well Cell Migration Assay kit (Cell Biolabs Inc, CBA-106, San Diego, USA) was used in order to evaluate THP-1 monocyte migration through a polycarbonate 8-µm pore size membrane. A total of 5×10^4^ cells/well transfected with 40 nM of hsa-miR-125b mimic, 40 nM of scramble or non-transfected controls, were seeded in the upper chamber side with RPMI1640 culture media without supplementation. The lower chamber side was filled with RPMI1640 media with the appropriate chemoattractant, MCP-1 or CCL21, using two different concentrations, 0.2 mg/mL or 40 mg/mL. After incubation of 24 h at 37 °C, the lower chamber migrated-cells were resuspended and dyed with CyQuant GR Dye provided with the kit and fluorescence at 460/560 nm was measured in the FLUOstar OPTIMA Plate Reader (BMG LabTech). The mean experimental values were normalized to control mean values.

### 2.11 Patients

This study employed samples from ascendant aortas obtained in the surgery scenario of coronary artery disease (CAD) patients and non-CAD controls. Samples and data from patients included in this study were provided by the Biobank HUB-ICO-IDIBELL (PT17/0015/0024), which is integrated in the Spanish National Biobanks Network. The study was approved by the ethics committee of Bellvitge University Hospital (PR149/14, approved on May 19th, 2016) and all participants provided their written informed consent. The demographic characteristics have been previously published^30^.

### 2.12 Statistical analysis

Data from at least three independent experiments or a single representative experiment that was performed in triplicate (unless otherwise noted) were presented as the mean ± standard deviation (SD). The Shapiro-Wilk and D’Agostino & Pearson normality tests were performed to identify normal distribution of variables. For the data that passed normality tests the statistical comparisons were made using an unpaired t-test or the two-way ANOVA with a Sidak’s post hoc test for multiple test correction (p ≤ 0.05). Data that did not pass the normality tests, comparisons for three or more categorical independent groups were made using non-parametric Kruskal-Wallis test corrected with Dunn’s post-hoc method (p ≤ 0.05) or Mann-Whitney for two groups (p ≤ 0.05). Significance was accepted at the level of p ≤ 0.05. Statistical analysis and normality tests were all performed using GraphPad Prism 8.0 for Windows (GraphPad software, San Diego, USA).

## 3. Results

### 3.1 MiR-125b inhibition has no impact on cholesterol levels or body weight but attenuates systolic blood pressure (SBP)

The body weights before and after 4 consecutive weeks of treatment, did not differ with regard to the groups of treatment (Table 1). Differences regarding lipid metabolism were measured in plasma samples from each mice group in order to determine whether these parameters are closely related to atherosclerotic plaque development. Levels of HDL and LDL cholesterol did not differ between groups. Although triglycerides (TG) levels were slightly higher in the groups treated with Anti-125b and miR-125b with respect to the control, this parameter does not correlate with an increase in plaque lesion. As observed below, the group treated with Anti-125b presented lower plaque size.

**Table 1.**
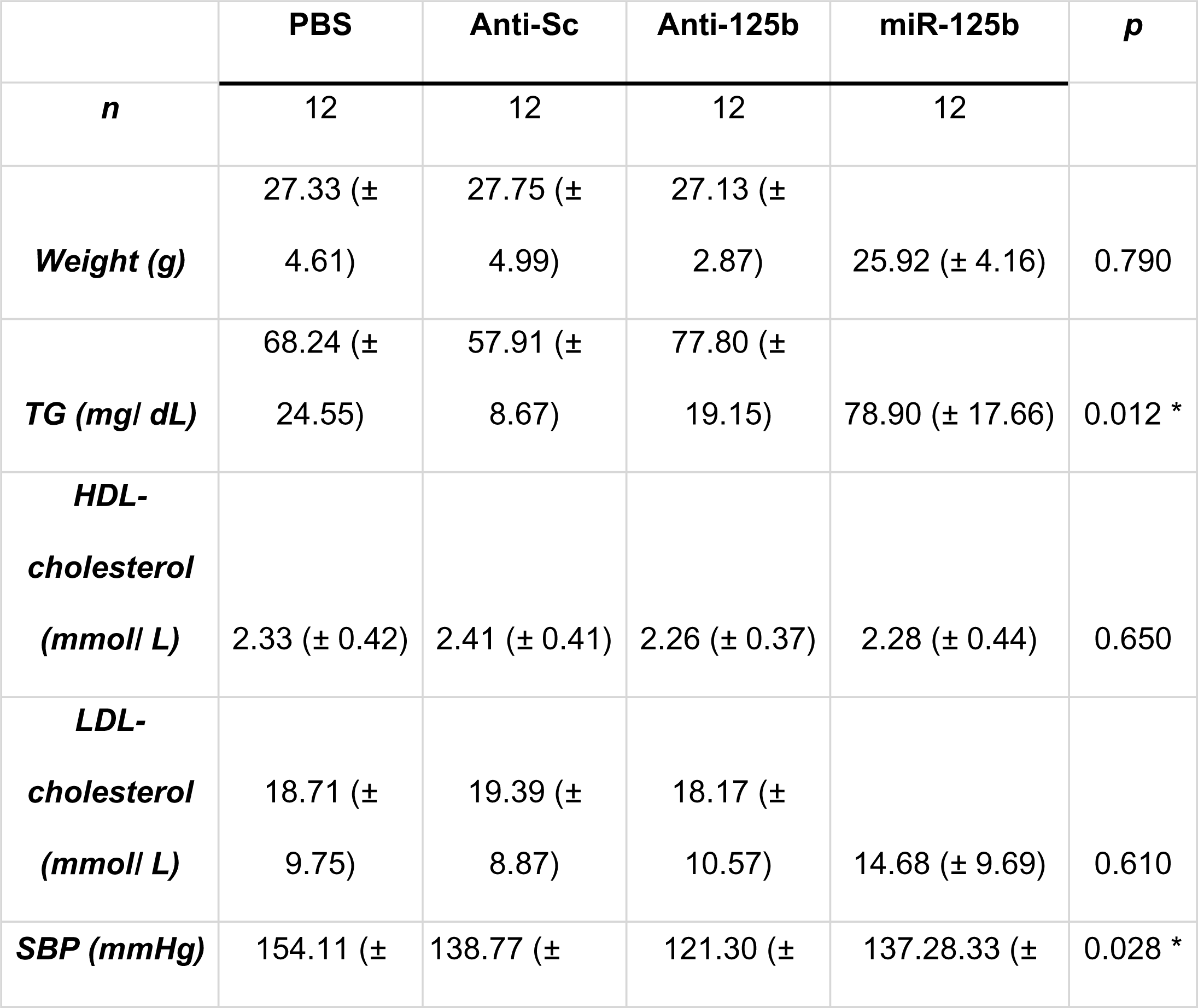
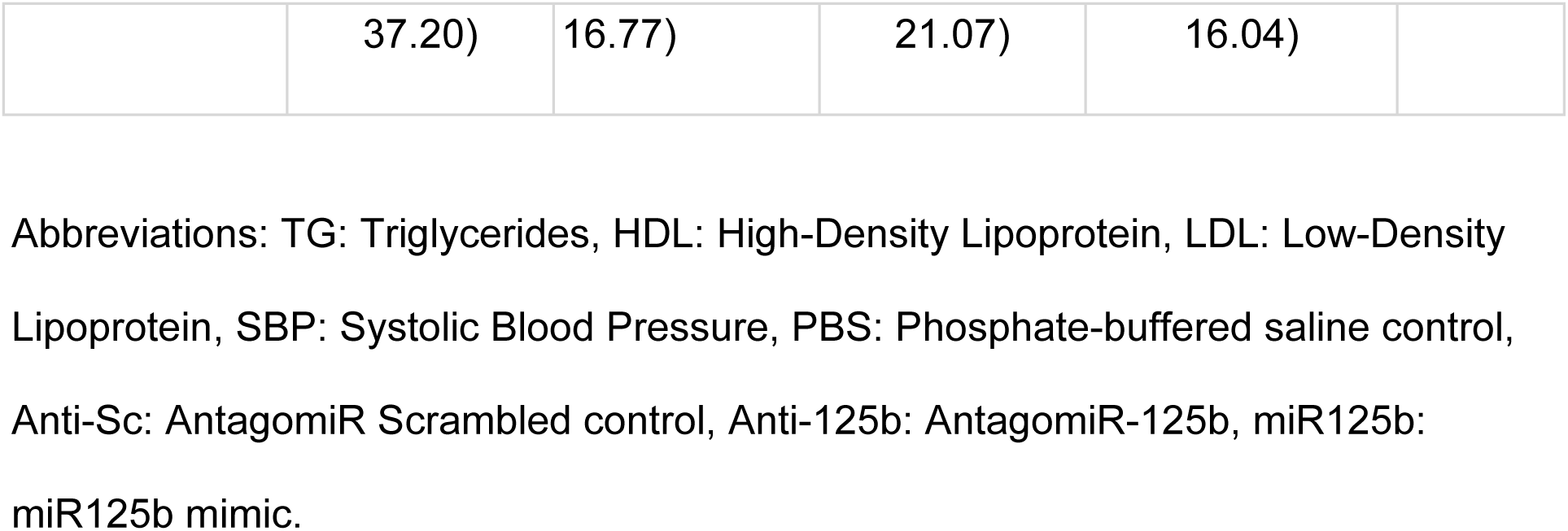
Plasma, blood pressure and weight parameters levels of High-Fat Rodent Diet - Treated ApoE^-/-^ mice. * p compared to Anti-Sc.

The non-invasive tail-cuff method revealed that systolic blood pressure of anti-miR125b-treated group was reduced when compared to PBS and Anti-Scramble control groups of ApoE^-/-^ mice maintained on High Fat Diet for 14 weeks (Table 1). Apart from recognizing miR-125b’s involvement in the regulation of the blood pressure system, as established by Chen et al. in 2021, further support for this understanding is derived from the research conducted by Nagpal et al. in 2016. Their findings indicate that inhibiting miR-125b effectively mitigates hypertension-induced cardiac fibrosis, which are in line with our results^31^.

### 3.2 Anti-miR125b treatment attenuates atherosclerosis progression

To test the hypothesis that miR-125b is implicated in atherosclerosis formation and progression in vivo, we designed an experiment using mice treated with mimics or inhibitors of miR-125b as it is summarized in experimental outline in Figure 1. We evaluated and quantified atherosclerotic plaque size and lipid neutral content on aortic sinus (Figure 2A and 2B) and in the whole aorta (Figure 2C) after 4 weeks of different treatments in the ApoE^-/-^ mice fed 14 weeks with HFD. The microRNA-125b inhibition treatment showed a notably reduction in the plaque size from the aortic sinus when compared to the Anti-Sc group (0.2907 ± 0.05mm^2^ in the anti-125b group vs. 0.4267 ± 0.07 mm^2^ in the Anti-Sc group; p < 0.002). Similar reduction of plaque lesion was obtained when compared total lesion area from the aortic arches of anti-miR125b mice group with Anti-Sc group (5.07 ± 5.02% in the Anti-125b group vs. 20.96 ± 7.68% in the Anti-Sc group; p < 0.001). Figure 2A and 2C showed in the right panels the quantification of lesion size by Trichrome’s method in aortic sinus and by ORO method in the en-face of the aortic arches respectively.

**Figure 2.**
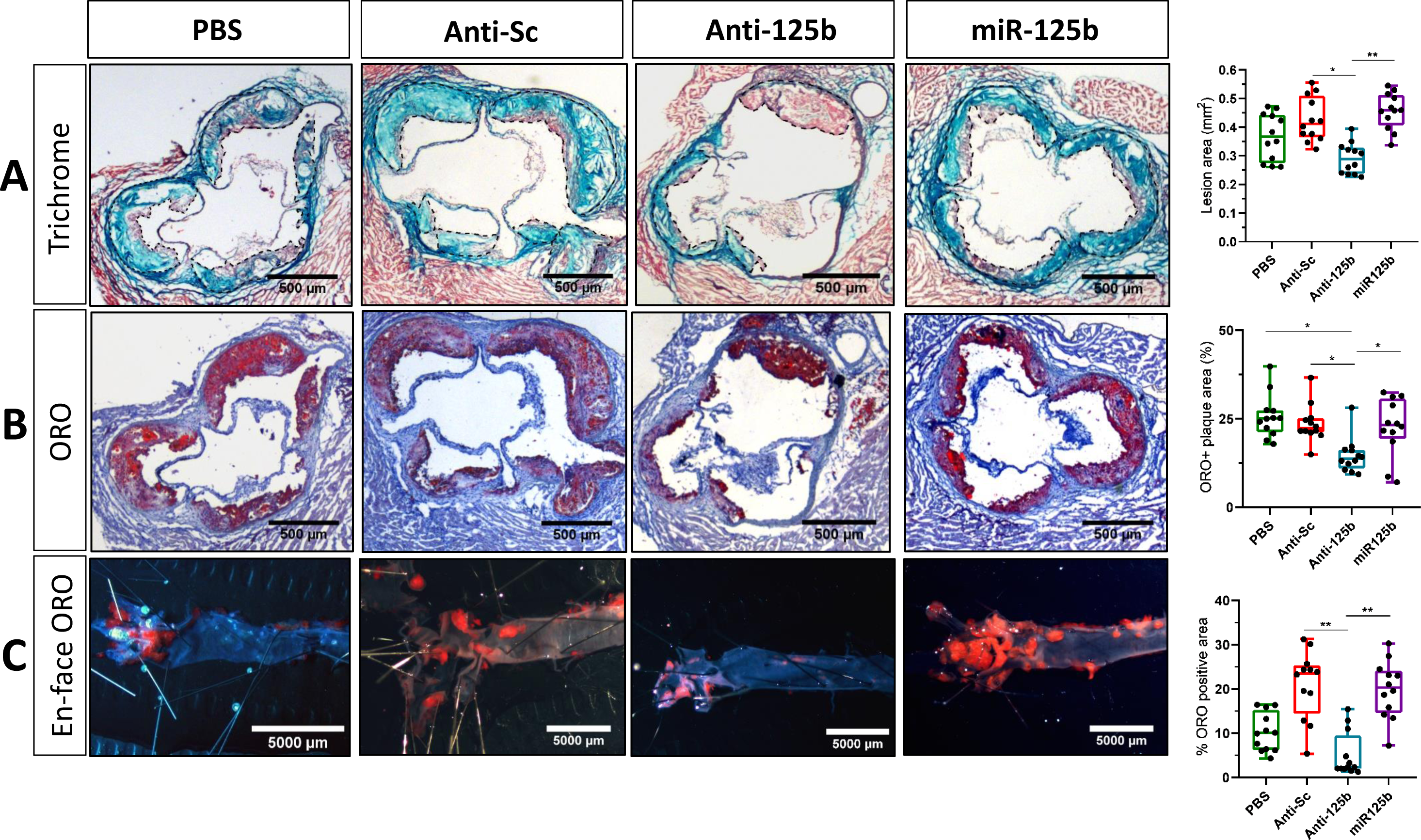
MicroRNA-125b inhibition treatment maintained for 4 weeks reduced atherosclerotic plaque lesion in ApoE^-/-^ mice. Representative histological analysis of cross sections of aortic sinus stained with Masson’s trichrome (A) or Oil -Red-O (B) from 22-week-old ApoE^-/-^ mice received intraperitoneal injections of PBS or 15mg/kg body weight antagomiR-Scramble (n=12), antagomiR-125b (n=12) or miR-125b mimic (n=12) twice a week for four weeks. In A the dashed lines delimit plaque lesion area and quantification of plaque size (lesion area) in mm^2^ is shown in the right panel as the mean±SD (n=12 mice per group; *P<0.05, **P<0.001). Quantification of neutral lipid accumulation in % within the plaque from the 4-treatment group is represented in the right panel as the mean±SD (n=12 mice per group; *P<0.05, **P<0.001) (B). (C) Representative en-face ORO staining of aortas from 22-week-old ApoE-/- mice aortas of corresponding 4 treatment groups. Scale bar, 5000 µm. Quantification of ORO positive areas in % is presented in the right panel as the mean mean±SD (n=12 mice per group; *P<0.05, **P<0.001). Data were analyzed by Kruskal Wallis nonparametric test with Dunn’s correction for multiple comparisons. ORO, Oil Red O; PBS, phosphate buffered saline; Anti-125b, Anti-microRNA 125b; Anti-Sc, Anti-microRNA Scramble.

Consistent with these results, we also observed a reduction in the neutral lipid content of the plaque within the aortic sinus from mice group treated with Anti-miR125b (14.58 ± 4.96% in the Anti-125b group vs. 23.67 ± 5.33% in the Anti-Sc; p < 0.001) as showed in the right panel of Figure 2B, suggesting that administration of anti-microRNA 125b reduces the plaque size and alters favorably the neutral lipid composition promoting an overall decrease in the atheroma burden. Accordingly we did not observe significative differences in plaque size of ApoE^-/-^ mice fed with HFD after administration during 4 weeks of exogenous miR-125b mimic compared to the Anti-Sc group neither in aortic sinus (0.456 ± 0.06 mm^2^ in the mimic-125b group vs. 0.426 ± 0.07 mm^2^ in the Anti-Sc; p > 0.99) nor in aortic arches (19.87 ± 6.4% in the mimic-125b group vs. 20.96 ± 7.68% in the Anti-Sc group; p= 0.96) or in neutral lipid content (22.64 ± 8.22% in the mimic-125b group vs. 23.67 ± 5.33% in the Anti-Sc group; p > 0.99).

### 3.3 MiR-125b inhibition reduces inflammation in Plaques

To assess whether miR-125b is associated with an inflammatory environment within the plaque of ApoE^-/-^ mice we used the histological cross-sections of aortic sinus roots adjacent to the ORO staining. Cellular plaque composition was evaluated by immunohistochemistry revealing that the relative content of F4/80 macrophages in the plaque lesions from aortic sinus was significantly decreased in ApoE^-/-^ mice treated with anti-miR125b (Figure 3A). Quantification of F4/80 macrophage population in Figure 3C showed less macrophage infiltration in anti-125b treated group compared to Anti-Sc group (0.145 ± 0.040% in the Anti-125b group vs. 0.386 ± 0.0574% in Anti-Sc; p<0.0001) or mimic miR-125b group (0.145 ± 0.0402% in the anti-125b group vs. 0.409 ± 0.0247% in the mimic miR-125b group; p<0.0001).

**Figure 3.**
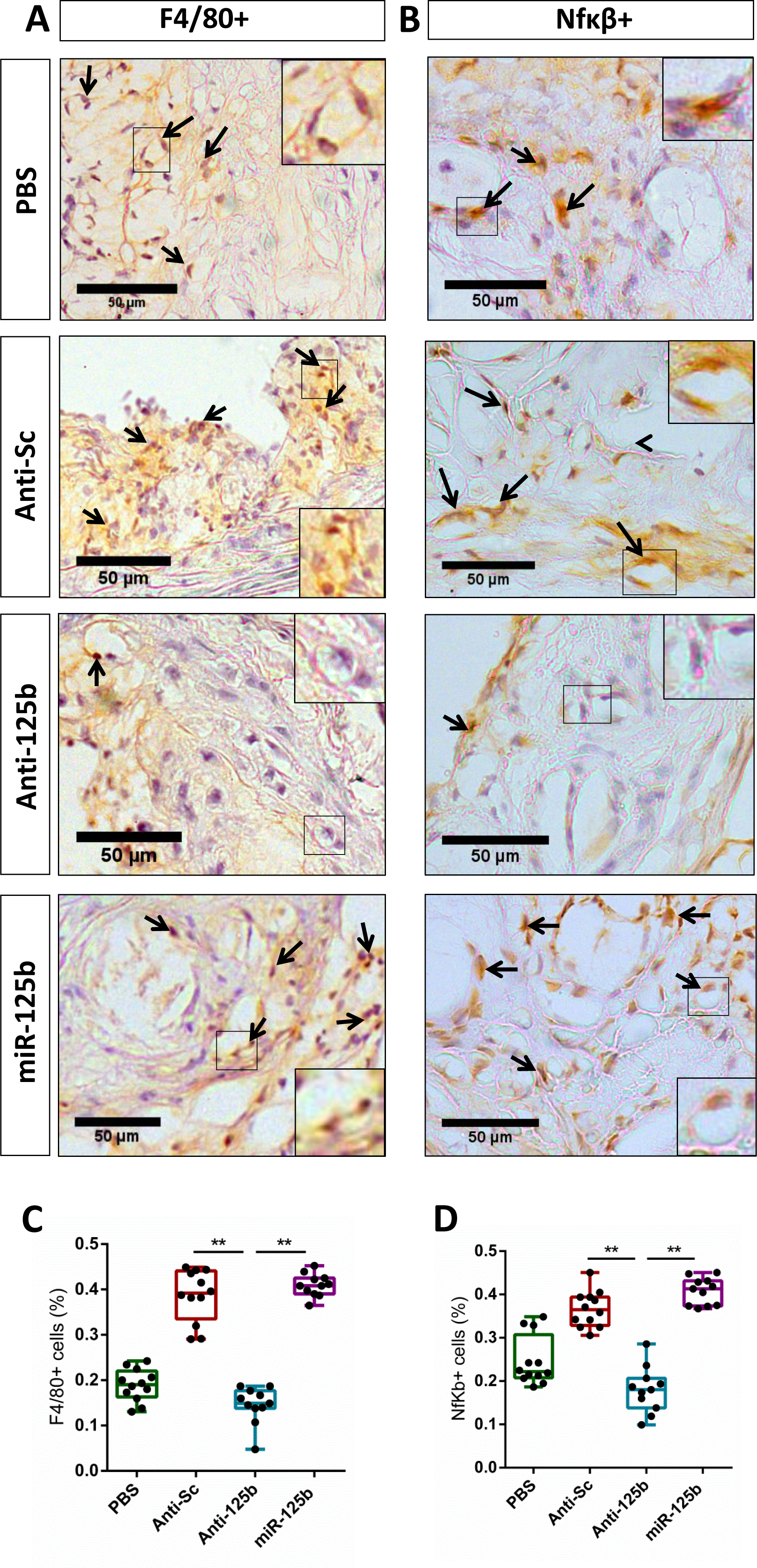
Anti-miR-125b treatment reduced macrophage content in atherosclerotic lesions from ApoE^-/-^ mice. (A) Representative immunofluorescence staining of macrophages (F4/80 positive) in cross-sections of the aortic roots from ApoE^-/-^ mice that received intraperitoneal injections of PBS or 15mg/kg body weight antagomiR-Scramble (n=12), antagomiR-125b (n=12) or miR-125b mimic (n=12) twice a week for four weeks. Scale bar, 50µm. (B) Representative immunofluorescence staining of NfκB positive cells in cross-sections of the aortic roots from ApoE^-/-^ mice injected during 4 weeks with PBS, Anti-miRNA Scramble 15 mg/kg, Anti-miRNA 125b 15 mg/kg or mimic miRNA-125b 15 mg/kg twice a week for four weeks. Scale bar, 50µm. The magnifications evidencing positive cells (or negative cells in the Anti-125b group) are shown in the insert (right quadrants of each immunostaining). Black arrows indicated examples of positive cells for F4/80 or NfκB. (C and D) Quantification of F4/80+ and NfκB+ cell content in % is shown in the lower panels expressed as mean±SD (n=12 in PBS and anti-miRNA Scramble, n=11 in Anti-miRNA 125b and miRNA-125b group; *P<0.05, **P<0.001 vs group treated with the Anti-miRNA Scramble). Data were analyzed by Kruskal-Wallis non-parametric test followed by the Dunn post-hoc correction test. Anti-125b, Anti-miRNA 125b; Anti-Sc, Anti-miRNA Scramble; NfκB, Nuclear factor kappa-light-chain-enhancer of activated B cells; F4/80, EGF-like module-containing mucin-like hormone receptor-like 1.

NF-κB transcription factor marker of inflammation was also assessed in the adjacent aortic sinus sections. In accordance with the F4/80 macrophages content, the ApoE^-/-^ mice treated with microRNA-125b inhibitor demonstrated a marked reduction of relative NF-κB^+^ cells in plaques (Figure 3B). Concordant with the in vivo histology’s observation, quantification of NF-κB+ cells (Figure 3D) showed a notable reduction in the anti-miR125b group when compared with Anti-Sc group (0.179 ± 0.0526% in the anti-125b group vs. 0.366 ± 0.0415% in the Anti-Sc; p=0.0002) or mimic miR-125b group (0.179 ± 0.0526% in the anti-125b group vs. 0.407 ± 0.0311% in the mimic miR-125b group; p<0.0001).

These findings suggest that anti-miR125b administration decreases macrophage content and reduces inflammatory environment in the plaques in aortic sinus from ApoE^-/-^ mice thus contributing in the mitigation of atherosclerosis.

### 3.4 MiR-125b regulates monocyte trafficking to plaques and downregulates monocytic CCR7 expression

In order to observe the implication of miR-125b in the molecular mechanisms of rolling and adhesion of monocytes to endothelial cells, the Vas on Chip adhesion assay was designed. We sought to investigate how shear stress conditions would affect transfected monocytic THP-1 cell adhesion to stimulated HAoEC.

Stimulated endothelial cells profoundly enhanced monocyte adhesion as we can observe in static conditions (Figure 4A). Although less attachment was observed in miR-125b transfected THP-1 cells, no significant differences were identified respect to the control group transfected with scramble (85.07 ± 26.77 cells vs. 114.8 ± 59.51 cells, respectively; p= 0.238) when we quantified the average cell attachment (Figure 4C). This scenario changed when we applied 12 dyn/cm^2^ high shear stress conditions, in which we observed less monocyte attachment as expected (Figure 4B), but this time we identified a remarkable decrease of THP-1 attachment in the miR-125b transfected group compared to the scramble group (15.07 ± 11.27 cells vs. 72.82 ± 46.50 cells, respectively; p<0.0001) as shown in analysis of the average cell attached in Figure 4D.

**Figure 4.**
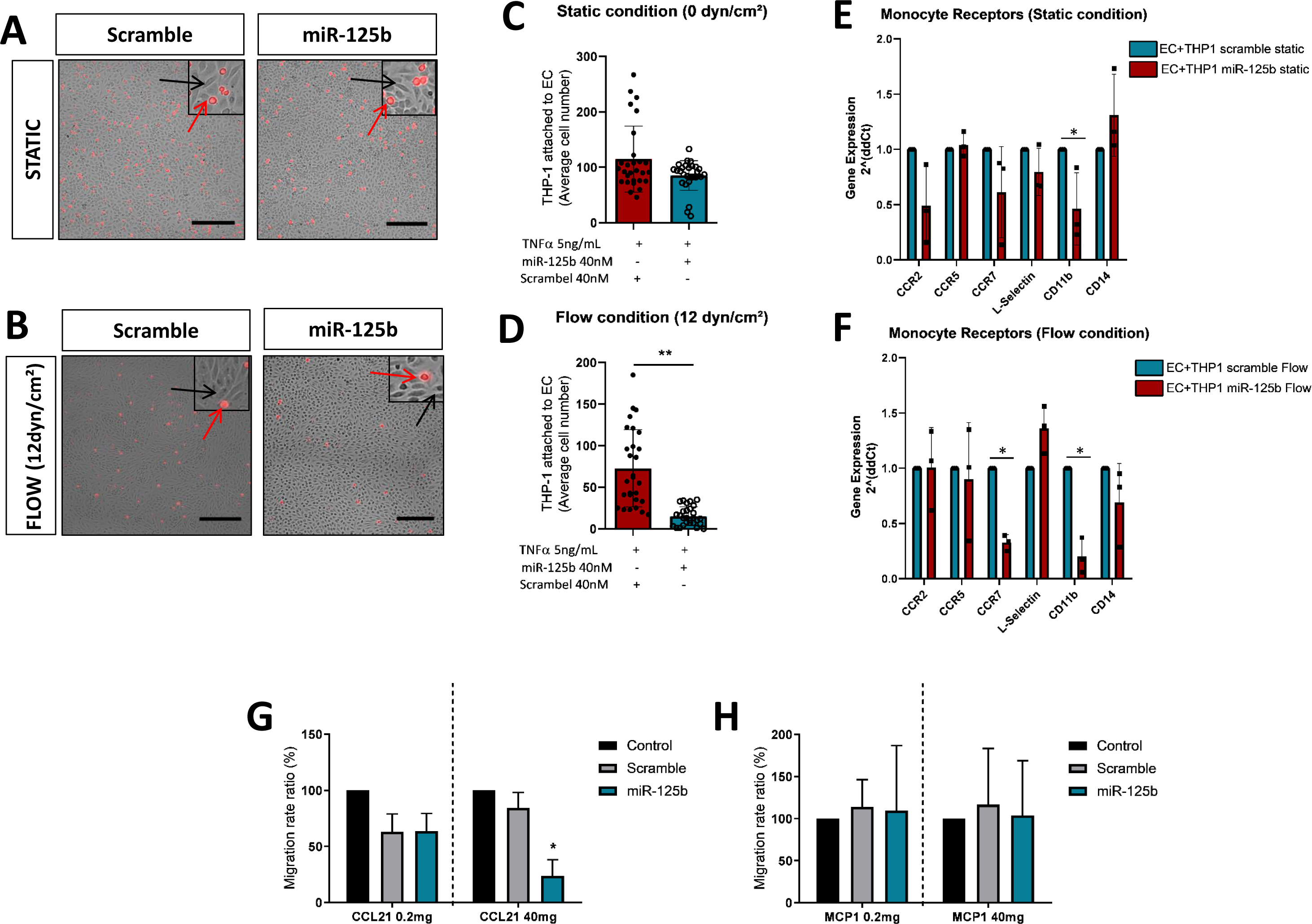
Monocyte adhesion and migration is regulated by miR-125b. Vas on a Chip assay (VoCh) was performed in order to assess the adhesion behavior of transfected monocytes to activated endothelial cells. (A) Representative confocal microscopy images of Calcein Red-labeled THP-1 monocytes attached over TNFα-treated HAoEC inside Leuer0.6 chambers after VoCh assay under static conditions (0 dyn/cm^2^) or (B) under flow conditions applying shear stress (12 dyn/cm^2^). The left columns showed adhesion of fluorescently labeled THP-1 transfected with miRNA Scramble and the right columns showed the adhesion of fluorescently labeled THP-1 transfected with miRNA-125b. Scale bar, 150µm. Each image showed brightfield channel exhibiting HAoEC (grey) and Fluorescent 590 nm red channel exhibiting THP-1 (red). The magnifications showed in the right upper quadrants from each image indicated examples of HAoEC (black arrows) or THP-1 (red arrows). (C) Quantification of monocyte adhesion on TNFα-treated HAoEC using microscopy under static conditions or under (D) flow conditions (12 dyn/cm^2^) expressed as the mean±SD of the average number of transfected THP-1 attached from 8 randomly selected fields (n=3 in each group; **P<0.001). Data were analyzed by Mann-Whitney non-parametric two-tailed test. (E) Quantification of monocyte receptors expression CCR2, CCR5, CCR7, L-Selectin, CD11b and CD14 in THP-1 using RT-qPCR after HAoEC activation with TNFα under static conditions or (F) after applying shear stress (12 dyn/cm^2^) and comparing miRNA Scramble-transfected THP-1 cells (blue columns) with miR-125b-transfected THP-1 cells (red columns) (n=3 in each group, *P<0.05). Data were analyzed by unpaired two-tailed t-test (G) Quantification of THP-1 monocyte migration rate from THP-1 monocytes alone (black columns), THP-1 transfected with miRNA Scramble (grey columns) or THP-1 transfected with miR-125b (blue columns) after 24 h incubation with CCL21 chemoattractant at low concentrations (left panel) and high concentrations (right panel) or (H) with control chemoattractant MCP-1 at low concentrations (left panel) and high concentrations (right panel) expressed as the mean±SD (n=3 in each group; *P<0.05). Data were analyzed by ordinary two-way ANOVA followed by Sidak’s multiple comparisons test. VoCh, Vas on Chip; THP-1, human leukemia monocytic cell line; HAoEC, Human aortic endothelial cells; TNFα, tumor necrosis factor α; CCR2, C-C chemokine receptor type 2; CCR5, C-C chemokine receptor type 5; CCR7, C-C chemokine receptor type 7; CD11b, cluster of differentiation molecule 11B; CD14, cluster of differentiation molecule 14; RT-qPCR, real-time quantitative polymerase chain reaction; CCL21, C-C chemokine ligand type 21.

When we analyzed by qPCR the expression of some monocyte markers related with migration in THP-1 after HAoEC activation with TNFα under static conditions we found that miR-125b downregulates the expression of CD11b/CD18 integrin, which enhances cell adhesion through its ligand ICAM-1 expressed in inflamed EC, compared to scramble transfected cells as shown in Figure 4E. Interestingly, under high shear stress the expression of CD11b/CD18 integrin and the receptor CCR7, implicated in the exit of macrophages from the plaques, were both decreased (Figure 4F). These data suggest that the trafficking of monocytes is altered by miR-125b.

To confirm the functional implication of CCR7 regulated by miR-125b in the egress of foam cells from the plaque we evaluate THP-1 monocyte migration. As shown in Figure 4G, the miR-125b transfected monocytes inhibited CCR7 cell migration induced by CCL21 ligand. In the same background, the miR-125b did not affect the MCP1-induced monocyte migration (Figure 4H) indicating that miRNA-125b acted particularly against CCR7 expression.

Because of the noteworthy impact of CCR7 in the miR-125b-transfected monocytes migration, we evaluated the expression of CCR7 receptor in THP-1 cells transfected with miRNA Scramble or miR-125b mimic, thereby providing evidence that overexpression of miR-125b results in a significant downregulation of CCR7 levels (Figure 5A). We hypothesized that miR-125b is a direct target of CCR7 gene. Thus, we demonstrate that miR-125b directly regulates CCR7 expression using a reporter plasmid construct (p_CCR7.WT) containing the 3’UTR region of CCR7 gene fused with a luciferase coding sequence which, when transfected by miR-125b, decreases luciferase activity (Figure 5B). The mutation of the seed region for miR-125b in the 3’-UTR of CCR7 gene (p_CCR7.Mut) rescued the luciferase activity (Figure 5B, 5C), evidencing that miR-125b is indeed a direct but poorly conserved target of the CCR7 gene.

**Figure 5.**
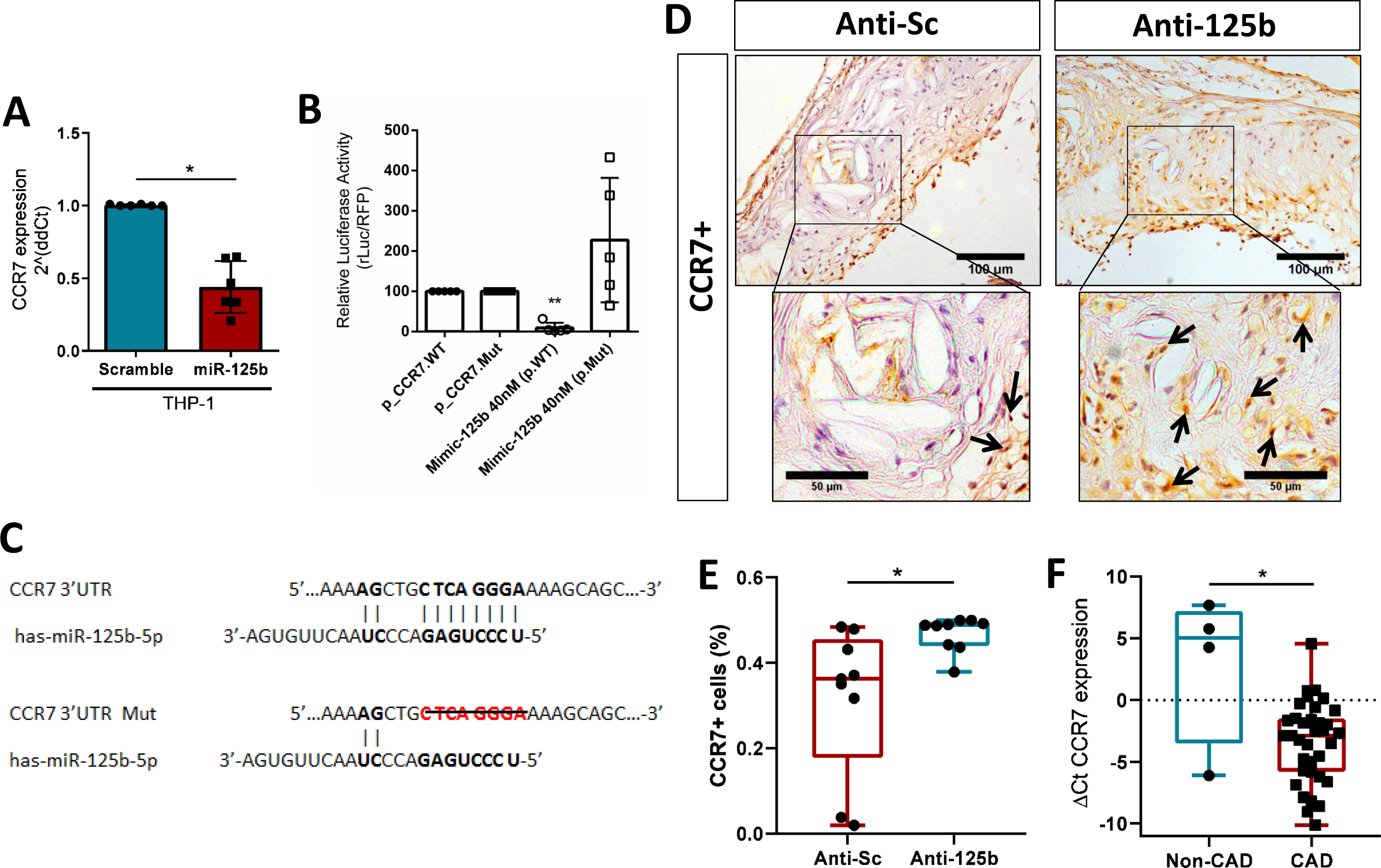
MiR-125b targets CCR7 and downregulates its expression in in vitro and in vivo. (A) Quantification of CCR7 expression in monocytic THP-1 cells 48 h after transfection with miRNA Scramble (blue column) or with mimic miR-125b (red column) using RT-qPCR (n=3 replicates; *P<0.05) and were analyzed using the non-parametric Mann-Whitney two-tailed test. (B) Luciferase activity assay 48 h after transfection of RAW264 murine macrophage cells that were co-transfected alone with pMirTarget vector containing the wild type 3-’UTR region of CCR7 gene (p_CCR7-WT) or pMirTarget vector containing the mutated 3’-UTR of CCR7 (p_CCR7.Mut) in the luciferase gene, or together with mimic miR-125b-5p. Luciferase values were normalized to the p_CCR7 control plasmids (the non-transfected with miRNA mimic). Data is expressed as the mean±SD (n=4 replicates in each group; *P<0.05 compared to non-transfected p_CCR7.WT) and were analyzed using the non-parametric Mann-Whitney two-tailed test. (C) Schematic representation of 3’-UTR region of CCR7 gene with the predicted binding site for miR-125b-5p (bold and linked). The sequence below shows the mutated CCR7 3’-UTR region with the deletion of eight nucleotides in the predicted binding site of miR-125b-5p (red bold, strikethrough). (D) Representative immunofluorescence staining of CCR7 positive cells in cross-sections of the aortic roots from ApoE^-/-^ mice injected during 4 weeks with Anti-miRNA Scramble 15 mg/kg and Anti-miRNA 125b 15 mg/kg. Scale bar, 100µm. The magnifications below evidenced positive cells (Black arrows indicated examples of CCR7 positivity). Scale bar, 50µm. (E) Quantification of CCR7 positive cell content in % is shown expressed as mean±SD (n=9 in each group; *P<0.05 vs group treated with the scramble antagomiR). Data were analyzed by using the non-parametric Mann-Whitney test. (F) Quantification of CCR7 expression in the aortas of patients with CAD (n=35) vs those without CAD (n=4) using RT-qPCR. Data is expressed as the mean±SD and were analyzed using the non-parametric Mann-Whitney test. 3’-UTR, 3-Untranslated region; Sc, Scramble, WT, Wild Type; Mut, Mutated; CCR7, C-C chemokine receptor type 7; RT-qPCR, real-time quantitative polymerase chain reaction; CAD, Coronary artery disease.

### 3.5 CCR7 is downregulated in coronary plaques both in the ApoE^-/-^ mice and in patients

Consistent with these findings, we evaluated the CCR7 expression by immunohistochemistry in the atherogenic plaque area of the ApoE^-/-^ animals treated as in Figure 1. The group treated with miR-125b inhibitor presented higher CCR7 receptor expression compared with the scramble group (Figure 5D), thus demonstrating that downregulating miR-125b, the CCR7 positive cells involved in macrophage migration are increased in the plaques from aortic sinus areas. Concretely, figure 5E showed a significant increase of macrophage infiltration in Anti-125b treated group compared to Anti-Sc group (0.468 ± 0.041% vs. 0.317 ± 0.173% respectively; P=0.0019). These results confirm that at the in vivo level, miR-125b also inhibits the CCR7 receptor and therefore, this microRNA can be a potential target to recover CCR7 in macrophages from atheroma plaques. Interestingly, these results were confirmed in human atherosclerotic lesions. As shown in Figure 5F, the cohort of patients with CAD that developed atherosclerotic plaques with an up-regulation of miR-125b^28^, also presented a reduction in CCR7 expression as compared with the non-CAD patients without atherogenic lesions.

## 4. Discussion

In this study, we demonstrated, for the first time to our knowledge, that therapeutic silencing of miR-125b in ApoE^-/-^ mice protects against atherosclerosis progression without significantly affecting circulating lipoprotein cholesterol. Mechanistically, we found that miR-125b regulates monocyte trafficking to plaques and downregulates monocytic CCR7 expression. Finally, we confirmed our results in a cohort of patients.

The microRNA miR-125b is mainly expressed in vascular smooth muscle cells (VSMCs) and macrophages, and its expression is upregulated under proatherogenic and proinflammatory conditions, such as the activation through the CD40-NF-κB pathway^27^. In addition, we demonstrated that miR-125b downregulates SR-BI and reverse cholesterol transport which contributes to foam cell transformation^28^. Here we have shown that suppression of miR-125b causes a reduction in the neutral lipid content of plaques. In addition, we observed a reduction in the macrophage cell content of aortic sinus plaques from mice treated with Anti-miR125b, supporting that atherogenesis is an inflammatory process. In this process, NF-κB activation is a key mechanism that leads to the production of a wide range of cytokines acting as key regulators in apoptosis, production of a proinflammatory milieu and plaque development^32,33^. In our study the immunohistochemistry analysis of the plaque revealed that relative content of F4/80+ macrophages along with NF-κB+ cells, were significantly decreased in the mice group treated with Anti-miR-125b. On the other hand, the group treated with a mimic miRNA-125b presented a higher number of cells with activated NF-κB+ within the plaque lesions.

Endothelial inflammation is likewise an important mechanism in monocyte recruitment within the subendothelial pro-inflamed wall. Recent in vitro evidences have demonstrated that the shear stress exerted on the atheroprone zone over the inflamed endothelium is also modulated by diverse miRNAs^34,35^. Moreover, in vivo inhibition of proatherogenic miR-92a, which is upregulated in atheroprone low SS areas, protects against endothelial dysfunction and ATH development^36^. Hence, using the Vas-on-Chip (VoC) technology we observed that under flow conditions the overexpression of miR-125b decreased monocyte adhesion to inflamed HAoEC. To deepen into the molecular mechanism we elucidate that, while CCR2 and CCR5, involved in adhesion and transmigration remained unaltered, there was a significant decrease in the expression of CD11b and, notably of CCR7. The integrin-subunit CD11b is also implicated during the rolling and adhesion process together with subunit CD18, forming the Mac-1 complex^8,37^. However, CD11b was not observed to be directly targeted and mediated by miR-125b as there was no similarity aligning their gene sequences. In contrast, we demonstrated that miR-125b directly targets and downregulates CCR7 gene. The 3’UTR of CCR7 gene contains a poorly but conserved seed region for miR-125b being a direct regulator of this receptor as we elucidated when the seed region was mutated in the luciferase assays. Furthermore, our in vitro functional studies of miR-125b overexpression were correlated with a decrease of CCR7 expression in THP-1 monocytic cells and demonstrate the impairment of monocyte migration in the presence of its ligand CCL21.

The CCR7 protein expression in the surface of macrophage and foam cells located within the plaque is essential to egress them into the lymph nodes, and the absence or downregulation has been shown to contribute to progression of ATH^8^. This insight is supported by the study of Trogan et al., which demonstrates that CCR7 was up-regulated in foam cells undergoing plaque regression in ApoE^-/-^ mice^10^. Other studies supporting this fact revealed that treatment with statins such as atorvastatin in ApoE-/- mice, allowed plaque regression depending on macrophage exit via the CCR7-signaling pathway^38^. Despite the accumulating evidence linking CCR7 implication to atherosclerosis attenuation, our study is the first to evaluate the therapeutic implication of miR-125b inhibition and the subsequent increase of CCR7 receptor levels within the plaque of aortic sinus from ApoE^-/-^ mice leading to an attenuation of atherosclerosis progression. Building upon our previous experiments we extended our investigations to assess CCR7 expression in aortas obtained from CAD patients. Specifically, we focused on individuals exhibiting up-regulation of miR-125b, where we observed a concurrent reduction in CCR7 expression. Atherosclerotic plaques are in part characterized by a dysregulation in CCR7 activity and expression which limit macrophage egress to lymph nodes exacerbating the inflammatory environment in plaque lesions thus impairing the resolution of inflammation. Here we reported that modulating the miR-125b/CCR7 axis may enhance the macrophage egress mediated by CCR7 promoting the resolution of inflammation, stabilization of plaques and limiting the atherosclerosis progression.

Further studies are required to determine how in vivo administration of miRNA mimics or inhibitors are delivered selectively into macrophages. The use of nanoparticles to target macrophages within aortic plaques that release the anti-miR125b will be a challenge for upcoming studies to better understand the miR-125b/CCR7 axis in diverse cell types. In this regard, our recent research has shown that incorporating an “eat-me” phagocytic signal cholesteryl-9-carboxynonanoate (9CCN) in the solid lipid nanoparticles (SLNs) together with the anti-miRNA target selectively the macrophages from aortic sinus of ApoE^-/-^ mice^39^.

To sum up, we present in this study the in vivo capacity of miR-125b inhibition-based therapy to reduce the plaque lesion size and the inflammatory plaque content. These improvements were, at least in part, mediated by the alteration in monocyte trafficking and CCR7 expression improving the exit of foam cells from the plaque. These results were confirmed in patients and have been useful in understanding the role of the miR-125b/CCR7 signaling in the resolution of atherosclerosis inflammation and its therapeutics implications. Thus, the inhibition of miR-125b alone or in combination with other miRNA targets implicated in the ATH events may represent a promising therapeutic approach for cardiovascular disease treatment.

## Data Availability

All experimental data referenced in this manuscript are available upon request. Researchers interested in accessing the data should contact to the corresponding author (MH).

## NON-STANDARD ABBREVIATIONS AND ACRONYMS

ATH: atherosclerosis;
CAD: Coronary Artery Disease;
CCL21: Chemokine C-C Ligand 21
CCR7: Chemokine C-C Receptor 7;
EC: Endothelial Cells;
HAoEC: Human Aortic Endothelial Cells;
HDL: High Density Lipoprotein;
MCP1: Monocyte Chemoattractant Protein 1;
MiRNA: micro-RNA;
OxLDL: Oxidized Low Density Lipoprotein;
ORO: Oil-Red-O;
Rt-qPCR: real-time quantitative Polymerase Chain Reaction;
SBP: Systolic Blood Pressure;
SLN: Solid Lipid Nanoparticles;
SS: Shear Stress;
SR-BI: Scavenger Receptor BI;
TG: Triglycerides;
TNFα: Tumor Necrosis Factor α;
VoC: Vas-on-Chip;
VSMC: Vascular Smooth Muscle cells;
3’UTR: 3’ Untranslated Region.
9CCN: cholesteryl-9-carboxynonanoate

## 5. Acknowledgments

We thank RICORS (RD21/0005/0001) and the CERCA Program - Generalitat de Catalunya for their institutional support. We want to acknowledge the patients and the Biobank HUB-ICO-IDIBELL (PT17/0015/0024) integrated in the Spanish National Biobanks Network for their collaboration.

## 8. Author contributions

M.H. and A.M. designed the study and wrote the whole manuscript. A.M., V.P., C.V. and L.M. contributed to the Vas on Chip design. E.N., A.M. and M.H. contributed to conceptualization. N.R., J.A. and J.C. contributed to the migration assays and the review of the manuscript.

